# Brain Ischemia in Alzheimer’s disease counteracts the Disruption of the Blood Brain Barrier: A Hypothesis Investigated with a Lumped Parameter Model

**DOI:** 10.1101/2024.12.18.24319268

**Authors:** Grant A Bateman, Alexander R Bateman

**Author notes:** Correspondence: Dr Grant Bateman Department of Medical Imaging John Hunter Hospital Locked Bag 1 Newcastle Region Mail Center 2310 Australia.

## Abstract

**Background:** In normal pressure hydrocephalus (NPH) there is blood brain barrier (BBB) disruption, which should increase the CSF formation rate (CSF_fr)_ and therefore the intracranial pressure (ICP). However, the ICP is normal in NPH. A lumped parameter study suggested that the CSF_fr_ could be reduced in this condition if the BBB disruption was moderated by a reduction in the capillary transmural pressure (TMP) secondary to arteriolar constriction. In early Alzheimer’s disease (AD), there is BBB disruption, reduced ICP and global ischemia. This raises the possibility that the same physiology may be occurring in AD as occurs in NPH.

**Objective:** This hypothesis can be analyzed further using a lumped parameter hydrodynamic model we have developed.

**Methods:** A lumped parameter model previously used to describe the hydrodynamics of NPH was modified to investigate the effects of changes in CSF pressure and blood flow in patients with mild cognitive impairment (MCI) and AD.

**Results:** The model indicates the capillary TMP is normal in MCI but decreases as AD progresses. Removing CSF in AD patients during a tap test initially increases the capillary TMP. The brain in AD responds to a tap test by increasing its level of ischemia and this reduces the capillary TMP.

**Conclusions:** A hypothesis is put forward that the BBB disruption in AD is partially mitigated by the brain making itself ischemic. Modelling gives support to this hypothesis. The model can explain the development of ischemic neuronal loss and amyloid accumulation secondary to glymphatic flow disruption as AD progresses.

## Introduction

Alzheimer’s disease (AD) is the commonest cause of dementia in the elderly population. It is characterized by brain atrophy and gradual cognitive decline, which correlates with a loss of neuronal synapses and cell death.^1^ The exact pathophysiology of this disease remains difficult to discern. There is evidence to suggest that blood brain barrier (BBB) disruption occurs early in AD. In a study of mild cognitive impairment (MCI) due to AD and early AD using an MRI gadolinium contrast protocol, there was global leakage of contrast, with a higher volume fraction of leaking brain tissue in the cortex, deep grey matter and normal appearing white matter correlating with disease stage.^2^ In both MCI and early AD, the mini mental state examination (MMSE) decreases significantly with increasing leakage of the BBB in the grey matter.^2^ The BBB protects the brain from blood derived toxic molecules, cells and microorganisms and also regulates the transport of nutrients and the clearance of metabolic end products and endogenous neurotoxins.^3^ The fact that the BBB breakdown occurs early in individuals with MCI and AD,^4^ even preceding hippocampal degeneration,^5^ suggests it may be an initiating event. In acute brain insults such as cerebritis, there is the production of vasogenic edema.^6^ Vasogenic edema is due to BBB disruption and results in extravasation of fluid and intravascular proteins such as albumin into the cerebral parenchyma.^6^ Excessive accumulation of this fluid evokes an increase in the intracranial pressure (ICP) because there is continuity between the interstitial and the subarachnoid spaces.^6^ Therefore, BBB breakdown in AD would be expected to increase the ICP. However, the opposite occurs, with a reduction in the ICP in AD over time. In a large study measuring the ICP in MCI and AD, there was a significant linear correlation between the reduction in the MMSE and a reduced ICP across the whole cohort.^7^ Thus, the progressive reduction in the ICP, despite the progressive opening of the BBB in AD, appears to be an enigma.

There is a high correlation between the pathology of AD and normal pressure hydrocephalus (NPH) found at brain biopsy performed whilst inserting a shunt to treat NPH.^8^ The syndrome of NPH was first described by Adams et al. almost 60 years ago, in patients with a classical clinical triad of ataxia, incontinence and dementia.^9^ These patients were found to have dilated ventricles but a normal cerebrospinal fluid (CSF) pressure.^9^ The ICP is normal in NPH despite there being a significant disruption of the BBB.^10^ As discussed, this suggests an anomaly in NPH pathophysiology similar to AD. Thus, there may be some pathophysiological overlap between NPH and AD. A lumped parameter hydrodynamic study of the brain in NPH performed by the current authors was performed to investigate this anomaly. This study suggested that the expected increased ICP in NPH could be moderated if the capillary pressure were reduced by arterial constriction leading to cerebral ischemia.^11^ Interestingly, a reduced cerebral blood flow (CBF) is also found to be a component of AD. A study by one of the current authors, showed the mean arterial pressure to be the same in a cohort of AD patients as compared to aged match controls. Using 2D phase contrast MR flow quantification, the total arterial inflow in the AD patients was measured to be 18% lower than the controls. Dividing the total blood flow by the mean arterial pressure suggested the vascular resistance in AD was increased by 23% (p=0.02) compared to the controls.^12^ This indicated a probable significant arterial dysfunction in AD, provided the capillary or venous pressure were not affected. In a large study, decreased global CBF was associated with worse cognitive performance in AD and impairment in all cognitive domains.^13^ There is a linear reduction in both CBF and the MMSE in AD patients over time, with a strong relationship between the decrease in global CBF and cognition.^14^ The similarities between the breakdown in the BBB, the reduced or moderated ICP and evidence of cerebral ischemia between NPH and AD is striking. This suggests to us a possible hypothesis i.e., the brain is inducing ischemia within its parenchyma in AD as a way of moderating the effects of the BBB disruption in a way similar to NPH. This hypothesis could be investigated further using the lumped parameter model we have previously developed. Therefore, the purpose of the current study is to extend the original lumped parameter model, to incorporate the ICP and ischemia in AD to test the hypothesis that the brain is reducing its blood flow as a way of moderating the ICP and the effects of the disruption of the BBB.

## Materials and Methods

A detailed description of the model can be obtained from the original paper.^11^ A brief description is given to outline the methods used.

### Equations

Davson’s equation relates the intracranial pressure to the CSF formation rate, the CSF outflow resistance and the venous sinus pressure,^15^

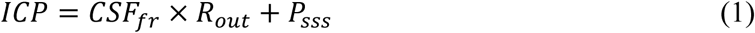

where ICP is the intracranial pressure, CSF_fr_ is the CSF formation rate, R_out_ is the CSF outflow resistance and P_sss_ is the pressure in the superior sagittal sinus. Next Ohms law for hydraulic circuits is required:

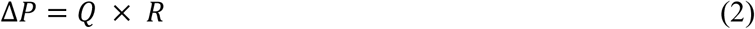

where ΔP is the pressure drop across a vascular segment, Q is the flow rate through the segment and R is the resistance. Resistances in series are additive so the following can be derived:

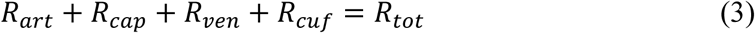

where R_art_ is the arterial segment resistance, R_cap_ is the resistance in the capillaries, R_ven_ is the venous resistance, R_cuf_ is the resistance of the venous outflow cuff and R_tot_ is the total resistance for the entire vascular system. Poiseuille’s equation calculates the pressure drop across each of these segments:

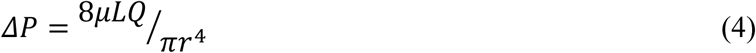

where ΔP is the pressure drop, µ is the viscosity, L is the vessel length, Q is the fluid flow rate, π is the circle proportionality constant and r is the radius. Substituting equation (2) into (4) and eliminating Q from both sides gives an equation for the resistance in each segment:

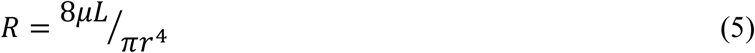

In this modeling study the viscosity, the length of each vessel segment, and π are constants, so it can be shown that a change in resistance for any segment depends only on a change in the vessel radius i.e.

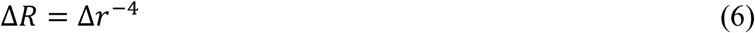

The volume of a vessel is given by the equation for a cylinder i.e.

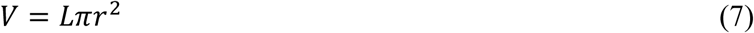

where V is the volume, L is the vessel length and r is the radius of the vessel. Given L and π are constants for any given segment, the change in volume is dependent on the change in radius i.e.

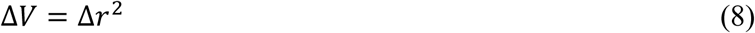

Substituting equation (8) into equation (6) gives

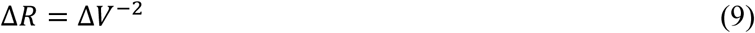

The next required equation relates the transmural pressure across a vessel to the vessel cross- sectional area:^16^

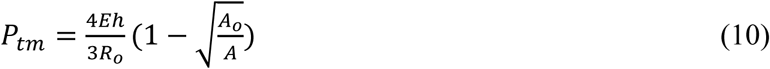

where P_tm_ is the transmural pressure across the vessel wall, E is the circumferential Young’s modulus of the vessel wall, h is the wall thickness, R_o_ is the radius in the stress free state, A_o_ is the area in the stress free state and A is the area following the applied transmural pressure. This equation was previously used to show that the volume of the venous outflow varies with the transmural pressure giving the equation:^11^

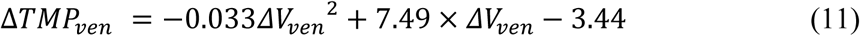

where ΔTMP_ven_ is the normalized increase in venous transmural pressure and ΔV_ven_ is the change in venous volume. The pressure within the venous outflow sinuses depends on the central venous pressure and the pressure drop across the venous sinuses to the level of the jugular bulbs. The normal central venous pressure is 5± 0.7 mmHg.^17^ Although the pressure drop across the venous outflow has been found to have a quadratic relationship with the CBF,^18^ the relationship of the portion of the graph between a normal flow rate and zero flow is almost linear. Thus, the venous sinus pressure can be found using the simplified equation:

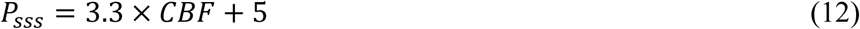

Where P_sss_ is the pressure in mmHg and CBF is the blood flow in L/min.

### Model input parameters

The input parameters are unchanged from the previous study^11^ and will only be briefly described as the details can be obtained from the original study. The brain size is 1500g. A CBF is 50 ml/100g/min.^19^ The cerebral blood arterial inflow is 750 ml/min. The arterial inflow pressure is 100 mmHg.^20^ The precapillary bed pressure is 32 mmHg.^21^ The end capillary pressure is 15 mmHg.^22^ The CSF pressure is 11.5 mmHg^23^ and the pressure gradient from the CSF to the superior sinus lumen is 4 mmHg,^24^ giving a sinus pressure of 7.5 mmHg.^25^ The transmural pressure of the subarachnoid cortical veins in primates is 2.5 mmHg.^26^ The pre-venous outflow cuff pressure is 14 mmHg by addition.

The total cerebral blood volume (CBV) is 51 ml.^11^ The arterial component of the CBV is 25% of the total^27^ or 12.8 ml. The capillaries make up 53% of the remaining volume,^28^ giving a total capillary blood volume of 20.3 ml and a total venous blood volume of 17.9 ml. The normal CSF formation rate is 0.40 ml/min.^29^

### Vessel responses to transmural pressure variations

The variations in the arterial resistance and volume in this model depend entirely on the arterial muscle tone and not the vessel transmural pressure. As the arterial pressure is always much higher than the ICP, the arterial transmural pressure will have no effect on the outcome of the current modelling study.

The capillary bed vessels do not actively alter their diameter,^30^ indicating they react purely to their transmural pressure. In a rat model, extreme hyperventilation decreased the PCO_2_ from 40 to 21.6 mmHg without affecting PO_2_ and the capillary size was not significantly different to the controls despite the expected arteriolar constriction.^31^ However, in the opposite case, in rats made extremely hypercapnic secondary to hypoventilation, the PCO_2_ increased to 95.6 mmHg but PO_2_ was normal, with the capillary diameter increasing by 20%, consistent with a 44% increase in volume compared to known control values.^31^ Thus, a moderate reduction in capillary trans mural pressure (TMP) does not change the capillary size but a maximal increase in TMP increases their volume by 44%. To simplify the current study, it is assumed the volume of the capillaries vary between normal and maximally dilated as a linear function of their transmural pressure. A previous study indicated an increase in capillary TMP from 12 to 37.9 mmHg would increase the capillary volume by 44% or a 1.7% increase in volume for each 1 mmHg pressure rise. Below a TMP of 12 mmHg, the volume is unchanged at 20.3 ml and above a TMP of 37.9, the elastic limit is reached and the volume is set to 29.2 ml.

Similar to the capillaries, the veins alter their size purely depending on their transmural pressures. In a previous modelling study,^11^ the function for the outflow vein dilatation was found to be summarized by equation (11).

At the distal end of the cortical veins, as they join the sinus wall, the outflow cuff segment resides. The collapse of this segment occurs physiologically and is passively modulated by the transmural pressure between the ICP and the sinus pressure, which is usually negative.^32^ The segment is very short, and as it is mostly under a state of collapse with physiological ICPs, the change in volume from this segment will be ignored in this model. This is despite acknowledging that the cuff is dilated in the tap test model. However, its resistance will be taken into consideration. In the previous study, four differing cuff transmural pressures resulted in 4 differing resistances.^11^ When these points were plotted, a straight line with R^2^ of 0.998 resulted, suggesting the cuff resistance varies as a linear function of the cuff TMP. Thus, giving equation 13:

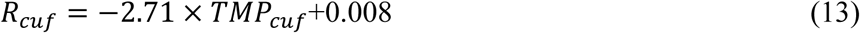

where R_cuf_ is the cuff resistance and TMP_cuf_ is the cuff transmural pressure.

The sagittal sinus pressure will vary with the blood flow using equation (12).

## Results

### Varying the Blood flow and ICP in AD

The original study findings in NPH suggested the brain elected to be ischemic, most likely to limit the capillary TMP and the CSF formation rate (CSF_fr_).^11^ We undertook an initial modelling study to see how the changes in CBF and ICP, known to occur in MCI and AD, would alter the capillary TMP to further study this effect. These initial modelling findings are summarized in Figure 1. The five vascular segments modelled are shown in fig 1a, each segment has a differing shade of grey. The segments from right to left are the arterial, the capillaries, the veins, outflow cuff and the sinus. The pressures obtained from the literature have been appended to the beginning and end of each vascular segment within the vessels in fig 1a. Given the arterial inflow volume passes through each segment sequentially, the resistance of each segment can be calculated using equation (2). These resistances are appended below the vessels in fig 1. The normal cerebral blood volume (CBV) values for each segment and the total CBV has been obtained from the literature and are shown below the resistances. The numbers above the vessel represent the transmural pressure gradients between the pressure at the beginning and end of each capacitance vessel segment and the ICP, and are obtained by subtraction. The extra figure above the capillary (in grey) is the mean TMP obtained by averaging the TMP before and after the capillaries. Placing the ICP, venous pressure and normal CSF_fr_ in equation 1 gives a normal R_out_ of 10 mmHg/ml/min. Figures 1b-e represent the effects of the differing alterations in CBF by varying the inflow resistance and ICP. In these figures, the darker grey segments represent the areas of increased resistance compared to the normal findings and the lighter grey represent reduced resistance.

**Fig. 1.**
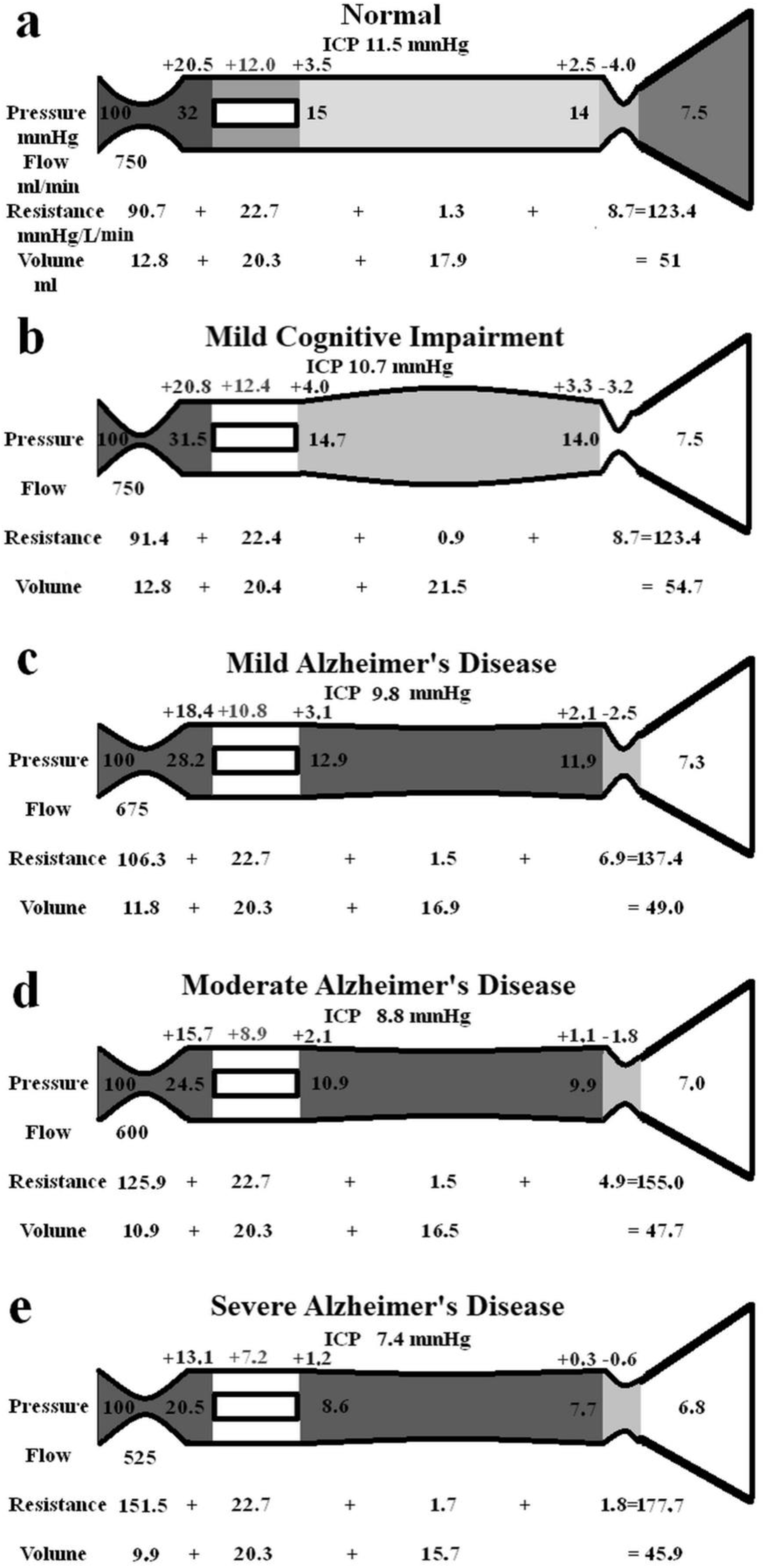
Results of modelling changes to blood flow in AD Fig. 1a depicts the normal findings. The segments from right to left are the arterial, capillary, veins, outflow cuff and the venous sinus. The vascular pressures are shown within the vessels. The numbers above the vessel are the transmural pressures at each site. The resistances and volumes for each segment are shown below the vessel. Note. ICP; intracranial pressure, mm; millimeters, mmHg; mmHg, millimeters of mercury, ml; millimeters. Fig. 1b shows the findings in MCI. The dark grey area indicating an increase in resistance in the arteries and the lighter grey decreased resistance in the veins compared to normal. Fig. 1c shows the findings in mild AD. The dark grey area shows a further increase in arterial resistance and some increase in venous resistance with the lighter area a reduction in cuff resistance. Fig. 1d shows the findings in moderate AD. These changes are essentially more pronounced than in mild AD. Fig. 1e shows the findings in severe AD. These changes have further progressed compared to earlier. Figure 1a has been reproduced from11 under a CC BY 4.0 commons licence

In figure 1b, the findings in MCI have been modelled. The arterial inflow is unchanged as per the literature. The ICP is reduced to 10.7 mmHg as per the literature. These pressure changes reduced the gradient pressure across the venous cuff and the resistance of this segment was reduced very slightly. The reduction in ICP dilated the veins due to the change in their TMP. There were no other significant changes. Placing the ICP, venous pressure and a normal CSF outflow resistance of 10 mmHg/ml/min into equation 1 suggests the CSF_fr_ is reduced in MCI to 0.32 ml/min, down from 0.4 ml/min.

Figure 1c models the findings in mild AD. The ICP and the CBF come from the literature. The venous sinus pressure is reduced, the cuff pressure gradient is reduced and the volume of the veins reduced. The effect was to reduce the capillary TMP below that for MCI. Placing the ICP, venous pressure and the normal R_out_ into equation 1 gave a CSF_fr_ of 0.25 ml/min, being a further reduction compared to MCI.

Figure 1d models moderate AD with further reductions in ICP and CBF. The effect is similar to, but more pronounced than the changes in mild AD, with the capillary TMP being further reduced. The calculated CSF_fr_ is also further reduced to 0.18 ml/min

Figure 1e models severe AD with further reductions in CBF and ICP. The capillary TMP is further reduced to 7.2 mmHg and the calculated CSF_fr_ is 0.06 ml/min. Note the total CBV has decreased by 10% compared to the normal figure.

### Performing a tap test in moderate AD

The effect of performing a tap test in moderate AD is modeled in figure 2. Figure 2a is the baseline moderate AD findings transposed from fig 1c for easier comparison.

**Fig. 2.**
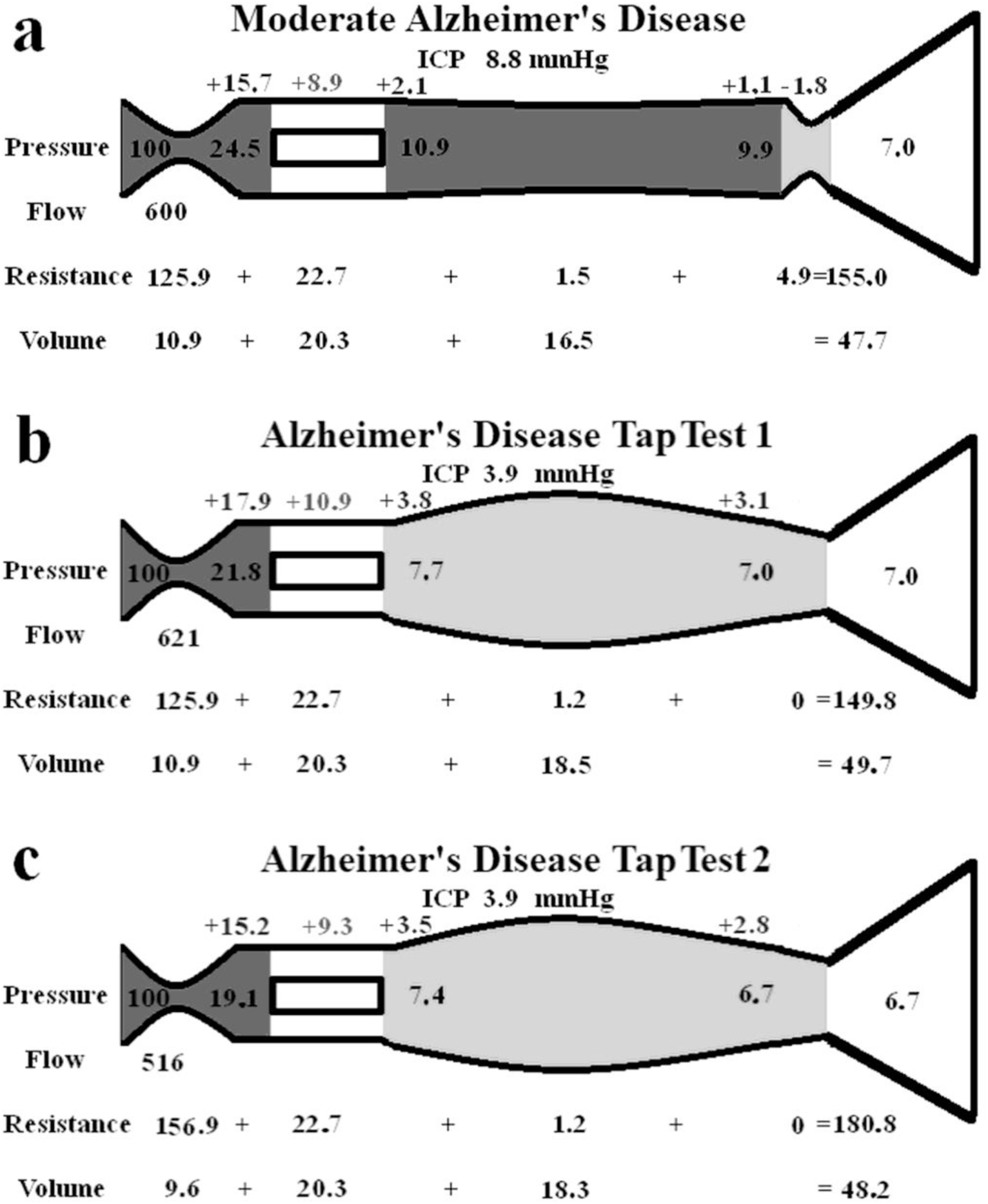
Modelling of changes secondary to a tap test in AD Fig. 2a shows the moderate AD findings reproduced from 1d for ease of comparison. Fig. 2b shows the immediate finding following a tap test. Note the veins dilate reducing the venous pressure and the flow increases slightly but the capillary TMP has increased. Fig. 2c shows the findings in a tap test 1 minute later as the CBF drops. This required an increase in the arterial resistance reducing the size of the arteries. The veins are dilated by almost the same amount meaning the total CBV is almost unaltered.

Figure 2b is the instantaneous effect of reducing the ICP by 56% as per the literature. The outflow cuff resistance is abolished due to the now positive pressure gradient across it. The unchanged arterial resistance allows for a minimal increase in CBF of 3.5%. There is a reduction in venous pressure but the ICP has dropped faster than this, so the capillary TMP increases by 22.5%. The lighter grey indicates dilatation of the veins and reduced resistance.

According to the literature the effect of a tap test is to induce a 10% reduction in CBF within several minutes. This effect has been modelled in fig 2c. The venous resistances are unchanged as they only depend on the transmural pressures which are minimally changed. A significant increase in arterial resistance is required, this reduced the capillary TMP by 14.7% compared to fig 1b because the precapillary pressure dropped and the venous sinus pressure dropped. The capillary TMP is reduced back towards the findings in fig1a. Note the CBV is not significantly different from 2a because the venous dilatation is matched by the arterial constriction.

## Discussion

We have hypothesized that the brain makes itself ischemic to reduce the effect of the BBB breakdown in AD. We wished to test this using a previously developed lumped parameter model. This model was originally verified by predicting the cerebral blood volume (CBV) changes which would occur at the limits of autoregulation, utilizing both an increase and decrease in perfusion pressure.^11^ The model accurately predicted the CBV changes found in the literature for both human and animal experiments.^11^ Similarly, we can check the accuracy of the current modelling in AD by comparing the predicted CBV with the literature. The model predicts a 10% reduction in CBV in severe AD. In a study of 16 patients with a MMSE averaging 16/30, the CBV was reduced by 11% in the cortex,^33^ suggesting the model is accurate enough for the current purposes.

### Cerebral blood flow in MCI and AD

A meta-analysis suggested no obvious changes in global CBF in MCI patients compared to controls.^34^ Therefore, we have not changed the CBF in our model for MCI. At first glance, a normal global CBF in MCI would appear to make our suggestion, that a reduced CBF is the earliest response to an opening of the BBB, unlikely. However, the perfusion changes in MCI are varied. In a recent MRI study, there was hypo-perfusion in the right middle frontal gyrus, right and left temporal gyrus and right middle temporal gyrus but hyper-perfusion in the right superior medial gyrus, left and right precuneus, left superior parietal lobule, right superior frontal gyrus, and right cerebellum.^35^ Thus, the high flow areas may mask the low flow areas in MCI at a global level. As previously discussed, there is a linear reduction in CBF over time in AD, with a close association between this reduction and the MMSE,^14^ suggesting the CBF reduction is closely related to the stage of the disease. In studies of mild AD with a MMSE of 26/30, the global CBF is reduced by between 6-10%.^36, 37^ We have chosen a figure of a 10% reduction in CBF for mild AD in our model. Studies of the CBF in moderate AD suggest a reduction of between 18-21%.^12, 38, 39^ Therefore, we have chosen a 20% reduction in CBF in moderate AD. There are few studies looking at severe AD and CBF. Tohgi et al. found a 25.7% reduction in the cortex and 17.8% in the white matter in severe AD.^33^ However, the global CBF can be reduced up to 42% in this disease.^40^ Given the linear response of the MMSE to the CBF, we elected to use a reduction of 30% in our model for severe AD. These reductions in CBF appear to occur despite there being no significant change in the arterial blood pressure.^12, 39^ Therefore, we have kept the inflow pressure normal in the model.

This progressive reduction in CBF in AD could be secondary to atrophy or reduced metabolic activity, however, published metabolic imaging suggests otherwise. Using Positron Emission Tomography and ^15^O labelled compounds, it is possible to measure the CBF in ml/100g/min, cerebral metabolic rate of oxygen (CMRO_2_) per 100g of tissue, oxygen extraction fraction (OEF) and CBV in ml/100g.^41^ CMRO_2_ measures the rate of aerobic glycolysis per 100g of tissue and the OEF is the fractional extraction of oxygen transferred from the capillary to the nerve cells.^41^ The cerebral metabolic rate of glucose (CMR_glu_) can be measured with a radio labelled glucose analogue and measures anaerobic glycolysis per 100g of tissue.^41^

When there is a fall in cerebral perfusion pressure, the first compensation measure the brain performs is to dilate the arterioles to decrease their resistance and maintain the CBF and CMRO_2_, this is stage 1._41_ Thus, the CBV should increase in stage 1. We note that the CBV actually progressively decreases in AD^33^ which is an anomaly. When the arterial vasodilatation is exhausted, the CBF will fall but the CMRO_2_ will initially be maintained by increasing the oxygen extraction with an increase in OEF, this is stage 2. If perfusion pressure falls further then infarction becomes a possibility.^41^ In 10 patients with AD judged to be moderate in severity, there were reductions in CBF, CMRO_2_ and CBV, with an increase in OEF compared to controls.^41^ There was a profound reduction in CMR_glu_, indicating reduced anaerobic glycolysis.^41^ The findings were described as misery perfusion. In this study the cerebrovascular reserve was assessed using 7% CO_2_ inhalation and hyperventilation to increase and decrease the CBF respectively.^41^ There was no difference in values between the AD patients and controls, indicating preserved cerebrovascular reserve.^41^ Another study found similar findings with large reductions in CBV, CBF, increased OEF and reduced CMRO_233_ and a third also indicated a 9% reduction in whole brain CMR_glu_ in early AD.^42^ This highlights a large discrepancy in AD. There is misery perfusion with metabolic derangement which is not just due to atrophy (the reduction occurs per 100g of tissue) but the arterioles are not fully dilated, as evidenced by the reduced CBV, and there is preserved cerebrovascular reserve. Therefore, the inference is that arterial constriction has caused the majority of the ischemia in AD and the arteries have elected not to dilate, even though they have the capacity to do so. In fact, the modelling in fig 2 suggests the arterioles actively react with a further constriction if the capillary TMP increases, suggesting that reducing the TMP may be a greater imperative than rectifying the metabolic insult.

A tap test is performed in NPH patients by removing 20-30 ml of CSF at lumbar puncture.

It has long been known that a tap test increases the CBF in NPH in those patients who improve clinically, but the CBV falls in those who do not improve clinically.^43^ In properly auto-regulated patients, increasing the cerebral perfusion pressure should not have an effect on the CBF. Even in idiopathic intracranial hypertension patients and those with a recent stroke, the CBF does not change with a tap test.^44, 45^ However, in one study where a tap test was performed on AD patients, the ICP was reduced by 56%, but the CBF fell by 14% within one minute. The CBV was not significantly changed.^44^ In another study, the tap test reduced the CBF by a further 10% on top of the 14% reduction in the baseline CBF in AD patients compared to controls.^46^ To study this effect, we reduced the ICP by 56% in fig 2b from the moderate AD baseline in fig 2a. The immediate effect would be a reduction in the venous outflow resistance and a moderate increase in CBF due to the improved perfusion pressure, fig 2b. However, the capillary TMP was increased by 22.5% in fig 2b. Following one minute, the CBF reduces, so we reduced the CBF by 10% of the normal flow rate in fig 2c to model this. This brought the capillary TMP down to be much closer to the original figure in fig 2a. Note the final CBV in fig 2c is only 1% larger than the baseline moderate AD value, despite the venous volume increasing by 11%. This is because the arterial volume decreased by 12% and this matched the venous increase. This CBV model prediction mirrors the findings of Meyer et al., in which the CBF fell but the CBV was unchanged in an AD at tap test.^45^ Again, this suggests our modelling is probably accurate enough for our current purposes. The interpretation is that the reduction in capillary TMP may be more important to the brain than the reduction in CBF that this requires.

### The blood brain barrier, CSF formation rate and the ICP

As already discussed, the ICP is reduced with disease progression in AD.^7^ In the study by Yang et al., the mean ICP in MCI was 10.7 mmHg, the patients with severe AD had an ICP of 7.4 mmHg and the average across all dementia patients (representing the level for moderate dementia) was 8.8 mmHg.^7^ We used these figures for our study. Given the linear relationship of the ICP with dementia grade, the value for mild dementia we used was taken to be half way between MCI and moderate dementia figure i.e., 9.8 mmHg.^7^

The intact BBB effectively excludes significant net fluid production within the brain parenchyma. The normal CSF_fr_ is 0.4 ml/min.^29^ Of this, 0.28 ml/min comes from the choroid plexus, 0.072 ml/min from the brain parenchyma and 0.048 ml/min from glucose metabolism.^47^ Therefore, the brain parenchyma produces some interstitial fluid, which becomes incorporated into the CSF production, at a rate of 0.0048 ml/100g/min. This compares to the remainder of the body where interstitial fluid production contributes to the lymph fluid. For the skin, the production rate is 0.24 ml/100g/min, for muscle it is 0.021 ml/100g/min^48^ and the liver is approximately 0.05 ml/100g/min^49^ or 2-3 orders of magnitude greater than fluid production within the brain. The fluid production can significantly increase in the brain parenchyma if there is opening of the BBB. Increased arterial pressure above the autoregulation cut-off gives cerebral hyper-perfusion, increased pressure within the capillaries and venules, disruption of the BBB, edema and a raised ICP.^30^ How much can a chronic disruption of the BBB increase the interstitial fluid production? In NPH there is BBB breakdown.^10, 50^ There is also a 14% reduction in the CBF in the cortex and a 40% reduction in the white matter.^51^ The capillary TMP in the cortex was modelled to be 9% above normal but in the white matter it was 33% below normal.^51^ It was argued an open BBB would make the cortex over-produce CSF but the white matter would absorb CSF and the balance would result in a normal ICP.^51^ Therefore, if this is true, we would expect there to be reversed flow in the aqueduct to accomplish this. Linstrom et al measured the CSF flow through the aqueduct in normal controls using phase contrast MRI and found it to be antegrade (coming out of the ventricles) at 0.18 ml/min but the flow was retrograde (going in) in 65% of patients with NPH who improved following shunt.^52^ These patients had 1.49 ml/min entering their ventricles.^52^ Given the grey matter makes up 65% of the brain, this would give a parenchymal interstitial fluid production rate of 0.15 m/100g/min, which is between the interstitial fluid production rate for the skin and liver as discussed previously. If the brain were not absorbing this fluid into the deep white matter at the same rate, then the 1.49 ml/min would be added to the 0.33 produced by the choroid plexus and glucose metabolism. Using Davson’s equation (1) the resultant CSF_fr_ of 1.82 ml/min, together with a normal R_out_ of 10 mmHg/ml/min and a normal sinus pressure of 7.5 mmHg, would give a resultant ICP of 25.7 mmHg in NPH, which is way above the normal range. Note, we used a normal R_out_ of 10 mmHg/ml/min in the moderate AD model and used Davson’s equation to calculate a net CSF_fr_ of 0.18 mls/min. This compares to Silverberg et al.’s finding of a net CSF_fr_ of 0.2 mls/min in moderate AD,^53^ suggesting our use of a normal R_out_ in AD is probably correct. The brain in NPH appears to make its deep white matter ischemic to allow the ICP to return to the normal range. In AD, unlike NPH, the maximal reduction in the capillary TMP is in the cortex and not the white matter^33^ and so excess CSF production would be moderated at its source. There is a progressive reduction in the ICP in MCI and with AD grade as shown in fig 1 and the net CSF_fr_ we estimated from Davson’s equation also reduces with MCI and with AD grade. This suggests that the progressive reduction in the capillary TMP we have estimated could be both correcting the over production of CSF and also reducing the net CSF production as well. Alternatively, the choroid plexus net CSF production may be being progressively down regulated secondary to the dysregulation of its protein synthesis.^54^ It is suggested altered gene expression within the choroid plexus produces down regulation of CSF production in AD.^55^ In MCI, the variable hyper-perfusion of brain regions with an intact BBB may be masking the effect of hypo-perfusion in areas with a deficient BBB in this global CBF study. However, the reduction in ICP in MCI suggests the hypo-perfused areas may be dominating with regards to a reduction in the CSF_fr_.

The possibility that the BBB disruption is being mitigated in the cortex could explain an otherwise difficult problem. The capillaries in AD have pathological changes of coiling and beading, as well as basement membrane disruption indicating BBB disruption.^56–58^ The ratio of the albumin concentration within the CSF to the serum (Qalb) is thought to represent a measure of BBB breakdown. The cut off for this metric is generally accepted to be 8, with a high number representing increased BBB breakdown.^59^ In AD the Qalb was measured to be 7.8± 4.8 in those with the highest quartile of ICP (early disease), 5.0± 1.5 in the next, 6.3± 2.7 in the next and 12.0± 12.7 in the lowest (severe disease).^7^ Therefore, according to the Qalb ratio, the BBB disruption only becomes a problem in severe AD and not early on (unlike all of the other evidence as presented). However, if the leakage of this protein were being opposed at the site of its production by a reduction in the capillary TMP (opposing the diffusion of the protein by reversing the CSF flow), then this unexpected result could be explained.

### Why is the blood brain barrier disrupted in AD?

The vascular hypothesis of AD was first suggested by de la Torre and Mussivand in 1993 and was a significant departure from the previous amyloid first paradigm.^60^ One of the current authors in 2004 extended the vascular hypothesis to propose that normal aging and senile dementia may be manifestations of a breakdown in arterial pulsation dampening, with either too large an arterial pulsation to be dampened, too small a compliance in the outflow pathways to allow dampening, or a combination of both.^61^ It was suggested that a form of pulse-wave encephalopathy could ensue if this dampening failed.^61^ This hypothesis was tested in a pilot study of 12 patients with mild to moderate AD, compared to 12 age matched controls. The mean arterial pressure was the same as the controls but the pulse pressure in AD was 9% higher. Using 2D phase contrast MR flow quantification, the arterial pulse volume within the arterial inflow flow was found to be 13.5% less than controls. Dividing the arterial pulse volume by the arterial pulse pressure suggested the compliance of the arterial tree in AD was reduced by 20% (p=0.05).^12^ Thus, in AD there is stiffening of the arterial tree which may increase the pulse pressure within the capillary bed due to a reduction in the dampening of this pressure from the reduced arterial compliance.^12^ The arterial inflow pulse pressure increases with aging, because the large elastic and muscular arteries become stiffer and the pulse pressure within the vascular system increases.^62^ The cerebral vessels also become stiffer with age.^62^ The increased pulse pressure is correlated with cognitive loss and neurodegeneration.^62^ A mouse model of carotid stiffness showed that blood brain barrier disruption occurred as a direct consequence of an increased pulse pressure.^63^ This represents the first part of the two-hit hypothesis in AD. According to the two-hit hypothesis, damage to the brain’s microcirculation occurs from aging and vascular risk factors such as hypertension, cerebrovascular disease, diabetes and hyperlipidaemia which lead to BBB dysfunction.^64^ This leads to the second hit i.e., increased amyloid β accumulation and impaired clearance.^64^

### The cause of the second hit

We have suggested that the initiating event in AD is breakdown of the BBB secondary to an elevated pulse pressure. The BBB disruption would be expected to increase the risk of toxic chemicals passing into the brain as well as the passage of bacteria and viruses. We suggest the brain mitigates this effect by making itself ischemic to reduce the leakage of these toxins. Unfortunately, this strategy is not without consequences. The prolonged ischemia will eventually lead to loss of synapses and whole neurones due to the accumulating ischemic damage.

The other problem is the accumulation of amyloid-β precursor protein components within the brain. Low CSF concentrations of amyloid beta (Aβ) products in patients with AD correlates with a high brain deposition, suggesting decreased clearance of these toxins.^65^ In early AD, before patients are symptomatic, CSF Aβ is reduced but there is increased amyloid PET tracer retention.^66^ After a variable lag period, neuronal dysfunction and neurodegeneration develop.^66^ Weller noted Aβ appears to accumulate within the perivascular interstitial fluid drainage pathways of the brain.^67^ The modern understanding of the interstitial drainage pathways comes from the glymphatic hypothesis. It is thought that CSF passes into the arterial perivascular spaces to percolate through the brain parenchyma and exit via the venous perivascular spaces into the subarachnoid space and also the lymphatics within the dura.^68^ Glymphatic fluid flow can be measured using an MRI technique called DTI-ALPS (diffusion tensor imaging along the perivascular space). Using this technique, there was noted to be a reduction in glymphatic function in the preclinical and prodromal stages of AD.^69^ A lower ALPS also predicts amyloid deposition, neurodegeneration and clinical progression.^69^ The entrance of the CSF into the periarterial space is thought to require a volume change within the artery i.e., pulsatile blood flow. As we have noted, the volume change in the artery in AD is reduced due to a lower arterial compliance.^12^ Unfortunately, this will be exacerbated by the progressive arterial contraction we hypothesise to occur with AD with disease progression.

Contraction of the smooth muscle cells in the arteries increases the wall tension and reduces the vessel compliance regardless of the pressure or diameter of the vessel.^70^ Thus, arterial contraction, in an effort to combat the BBB leakage will decrease the arterial volume pulsation and, therefore, the glymphatic flow leading to accumulation of these toxic proteins within the brain.

### Limitations

There are many assumptions inherent in the lumped parameter modelling. Poiseuille’s equation requires flow through a thin, rigid, circular tube of a Newtonian fluid, without turbulence. To the degree that these assumptions hold, the findings would be accurate. However, despite its limitations, this equation is commonly used in modelling the vasculature in the literature.

Some of the data we required for this model is not available from human studies. In its absence, animal studies were utilised. This is exemplified by the data linking the dilatation of the capillaries to the changes in TMP, which was taken from rodent studies and the normal venous TMP, which was obtained from primate studies. We have no way of knowing if the animal data closely approximates human findings, so this is a limitation. Despite this, the model seems to correctly predict the CBV in both severe AD and following a tap test.

## Conclusions

The earliest changes in the patients destined to develop AD are vascular stiffening and increase pulse pressure with disruption of the BBB. The brain appears to react by constricting its arteries to reduce the leakage of interstitial fluid and protein in to the parenchyma. This may work for some time and mitigate the risk of BBB disruption. However, eventually ischemic damage and a reduction in glymphatic flow will result. The later increases amyloid accumulation leading to further damage.

## Author contributions

Conceptualisation GAB, ARB. Methodology GAB, ARB. Validation ARB. Writing original draft preparation GAB. Writing- Review and editing GAB ARB. All authors read and approved the final manuscript.

## Data Availability

All data produced in the present work are contained in the manuscript

## Acknowledgments

The authors have no acknowledgments to report.

## Availability of data and materials

All data utilised in this study is contained within the article.

## Statements and Declarations Ethical considerations

Not applicable.

## Informed Consent Statement

Not applicable. Consent to participate Not applicable

## Consent for publication

Not applicable

